# A simple stochastic theory of extinction shows rapid elimination of a Sars-like pandemic

**DOI:** 10.1101/2020.08.10.20171454

**Authors:** Bhavin S. Khatri

**Affiliations:** Department of Life Sciences, Imperial College London, Silwood Park, Ascot, SL5 7PY, United Kingdom; The Francis Crick Institute, 1 Midland Road, London, NW1 1AT, United Kingdom

## Abstract

The SARS-Cov-2 pandemic has seen the challenge of controlling novel zoonotic diseases that have high infection fatality rates, including a natural capacity for the evolution of variants that transmit more easily and evade immunity. In dealing with current and future similar pandemics, the question arises: what is the optimum strategy to control infections. Although a complex question, a key neglected component to appraise the elimination strategy is simple theory predicting the expected timescales of elimination. We use simple random walk and branching process theory to provide new insights on the process of elimination using non-pharmaceutical interventions. Our central achievement is a full theory of the distribution of extinction times — which we find is an extreme value Gumbel distribution — for any value of the reproductive number including some degree of population immunity. Overall, for the original SARS-Cov-2 variant our results predict rapid extinction — of order months — of an epidemic or pandemic if the reproductive number is kept to *R*_*e*_ *<* 0.5; in a counterfactual scenario with global adoption of an elimination strategy in June 2020, SARS-Cov-2 could have been eliminated world-wide by early January 2021. Looking to the future, our results would suggest that elimination using NPIs is a more optimal strategy to control — and potentially eradicate — a Sars-like pandemic, in its early stages before the evolution of variants with greater transmissibility.

## Introduction

The SARS-Cov-2 pandemic has revealed a complexity of the interplay between the dynamics of infections, the evolution of the virus and the policies that governments have adopted to mitigate or eliminate the epidemic. The predictions of the SIR model *(1, 2)* would seem inadequate when compared to the history of infections seen during the SARS-Cov-2 pandemic, which would have allowed infections to rise until herd immunity brings infections under control; instead when faced with the high burden of hospitalisation and mortality, the majority of governments decided on alternative strategies of mitigation or elimination, which to different degrees use non-pharmaceutical interventions (NPIs) to control infections *(3–6)*. However, in addition we have seen the evolution of new variants that are more transmissible and or gained the ability to evade immunity *(7–9)*. Ultimately, for the SARS-Cov-2 pandemic, vaccination has provided a largely effective prophylaxis, making the question of mitigation vs elimination less relevant, particularly in light of SARS-Cov-2 having evolved variants of far higher transmissibility, making the latter strategy beyond what is generally considered practicable.

However, should the globe face a similar Sars-like pandemic in the future the question remains: what lessons can be learnt and what is the optimal strategy *(10)*? Looking back, the mitigation strategy with SARS-Cov-2 has encountered a number of difficulties, despite an unprecedented pace of vaccine development: natural and vaccine induced antibody immunity is short-lived, and generally much less than a year *(11–13)*, vaccines do not sufficiently cut transmission *(14–20)* to achieve *R*_*e*_ *<* 1, vaccine roll-outs take significant time, vaccine hesitancy has been significant *(21, 22)* and finally persistent worldwide infections have led to the evolution of variants that are more transmissible and an ability to evade vaccine and natural antibody immunity *(23–25)*. On the other hand, a number of countries in the Asia-Pacific region adopted an elimination strategy that aimed to eliminate local infections though NPIs *(26, 27)*, which on many metrics has been seen to be successful *(28)*, but was ultimately abandoned by some countries in light of the advent of vaccines that prevent serious disease and the difficulty in using NPIs to control more transmissible variants *(29)*. However, despite this, the questions remains of whether elimination is a viable strategy: can infections be controlled at a small number, without requiring an “endless” lockdown, or can infections be completely eradicated.

Although, this is a complex question, using simple random walk theory and branching process analysis, we determine the full stochastic properties of the time to extinction. A stochastic approach is necessary as it accounts for the discreteness of individuals that will dominate the final stages of an epidemic, which cannot be captured by the deterministic SIR model. There has been considerable work done on understanding stochastic aspects of epidemics *(30, 31)* from the role of critical community sizes in diseases such as measles *(32, 33)*, stochastic phases in the establishment of epidemics *(34)*, to stochastic extinction. With regard stochastic extinction most results have been focussed on understanding the time to extinction either through the whole course of an epidemic *(35, 36)* or assuming a quasi-equilibrium has been reached through herd immunity in the population *(37)*. In essence, the situation faced by many countries at the beginning stages of the SARS-Cov-2 pandemic and before vaccination, has not been considered in previous modelling; at the early stages significant herd-immunity had not been achieved in the population, and there was the potential that NPIs alone could be used to reduce the reproductive number to less than 1 and give rise to extinction of an epidemic. Our branching process analysis draws on standard results on sub-critical branching processes that show the distribution of extinction times is an extreme value Gumbel distribution *(38, 39)*; our key contribution here is to show the range of applicability of these results for effective reproductive numbers less than a critical value 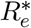 and, using a heuristic approach, further extend their range to the full domain of 0 *< R*_*e*_ *<* 1.

Using this new theory, our key practical finding is to show extinction/elimination can be achieved on the timescale of months, not years, and therefore there is a strong case that elimination should be the first defense against a new zoonosis like SARS-Cov-2 *(10, 28)*, before it has evolved greater transmissibility. Specifically, for an infectious disease like SARS-Cov-2, where infection durations are of order a week, reproductive numbers *R*_*e*_ *>* 0.6 give extinctions times which are long, of order many years, and dominated by the stochastic phase — on the other hand, extinction can be rapid with times much less than a year, or a few months, if restricted to *R*_*e*_ *<* 0.5. We show that if the globe adopted of an elimination strategy in June 2020, SARS-Cov-2 could have been eliminated world-wide by early January 2021. However, within the current context of a persistent epidemic that has evolved more transmissible variants and in the face of vaccines that only cut transmission moderately, this means that using NPIs to achieve a given *R*_*e*_ becomes proportionally more difficult. In light of this, we argue that elimination via NPIs is likely only optimal and feasible for early stages of a pandemic with moderate intrinsic transmissibility of the virus.

## Results

### SIR model with small fraction of infected individuals leads to non-changing *R*_*e*_

The SIR model divides the population of *N* individuals in a region into 3 classes of individuals: susceptible S (not infected and not immune to virus), I infected and R recovered (and immune, so cannot be re-infected). The model assumes a rate *β* of an infected individual infecting a susceptible individual (S + I → 2I), and a rate *γ* that an infected person recovers from illness and is assumed permanently immune/recovered. The model allows a simple characterisation of when infections will grow or decline: 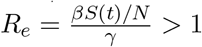, where *R*_*e*_ is dimensionless and is called the reproductive number (also commonly called *R*_*t*_) and will in general be time-dependent, as the number of susceptible individuals in a population change. *R*_*e*_ represents the average number of individuals an infected person infects through the duration of the infection *τ* = 1*/γ*. The classic course of an epidemic culminates in the herd immunity threshold being reached when sufficient number of individuals have become immune and *R*_*e*_ *<* 1 leads to a decline of infections to eventual extinction.

The use of non-pharmaceutical interventions (NPIs) provides an alternative means to reduce *R*_*e*_ by reducing the contact rate *β* between individuals to allow *R*_*e*_ *<* 1, without ever requiring herd immunity; this is the basis behind elimination strategies to control infectious diseases. As we show in the Supplementary Materials, as long as *R*_*e*_ is less than a critical value 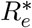 it is a good approximation to assume that the population of susceptible individuals *S(t)* = *S*_0_ is roughly constant and the reproductive number unchanging 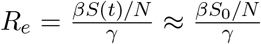; although each time an individual is infected, we loose exactly one susceptible the relative change of the susceptible pool is negligible, since the total number of susceptible individuals is very large. This critical value of the effective reproductive number is given by

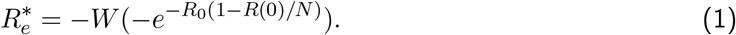

where *W* is the Lambert W-function. This regime corresponds to an initial value of *R*_*e*_ such that the decline is sufficiently rapid that the error due to ignoring the change in the susceptible pool is negligible. Calculating 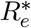 within the United Kingdom during summer 2020 infections were small *I*_0_ ≈ 3 × 10^4^ and assuming roughly 10% had recovered *(R*(0) = 0.1*N*), we find the constant *R*_*e*_ assumption to be good for *R*_*e*_ *<* 0.98, i.e. for all but *R*_*e*_ very close to 1, however, using numbers from January 2021, *I*_0_ ≈ 10^6^, and 15% recovered this requires *R*_*e*_ *<* 0.82.

This means for the case where only a small fraction of the population are ever currently infected, the SIR dynamics results in a single differential equation for 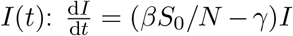. The solution to this is of course an exponential function: 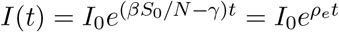, where *ρ*_*e*_ = *γ(R*_*e*_ − 1) is the effective growth rate for *R*_*e*_ *>* 1, and decay rate when *R*_*e*_ *<* 1, and *I*_0_ = *I*(0) the initial number of infected individuals. As we will see *ρ*_*e*_ more directly determines the dynamics of the extinction process than *R*_*e*_ or *γ* separately, and is in fact an easier quantity to determine (Supplementary Materials).

We are interested in understanding extinction of an epidemic and so from here on we define the rate *ρ*_*e*_ = *γ*(1 − *R*_*e*_) to be a positive quantity, making the assumption that *R*_*e*_ *<* 1. In this case we can make a simple deterministic prediction for the time to extinction, by calculating the time for the infected population to reach *I(t)* = 1:

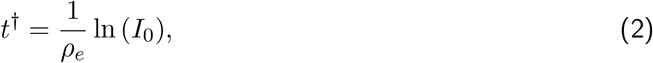

However, without understanding the stochasticity of the extinction process, it is difficult a priori to say anything about the goodness of this deterministic calculation. As we show below, using heuristic arguments, there is minimum number of infected individuals needed to overcome stochastic effects and confirm that this threshold also arises as a key determinant in the extinction time distribution in an exact branching process calculation.

### Stochastic extinction of an epidemic

The above analysis assumes deterministic dynamics with no discreteness – it ignores any randomness in the events that lead to changes in number of infected individuals. At the beginning of the epidemic, or towards the end, there are very small numbers of individuals infected, so these random events can have a large relative effect in how the virus spreads and need a stochastic treatment to analyse. We are interested in analysing the stochasticity of how the number of infected decreases when *R*_*e*_ *<* 1 and eventually gives rise to extinction, i.e. when there is exactly *I* = 0 individuals and the distribution of this time.

We first run stochastic continuous time simulations with Poisson distributed events (Gillespie or kinetic Monte Carlo simulations) *(40)* of the SIR model to confirm that the assumptions of a constant *R*_*e*_ due to a negligibly changing susceptible population of the previous section are accurate in the regime 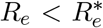. Fig.1a plots the decline in number of infected over time *I(t)*, where each of the trajectories from the Gillespie simulations is a grey curve, whilst the deterministic prediction (Eqn.2) is shown as the solid black line. We see that for *I(t)* ≫ 1 the stochastic trajectories are bisected by the deterministic prediction, indicating that the assumption of a constant *R*_*e*_ is a good one.

**Figure 1:**
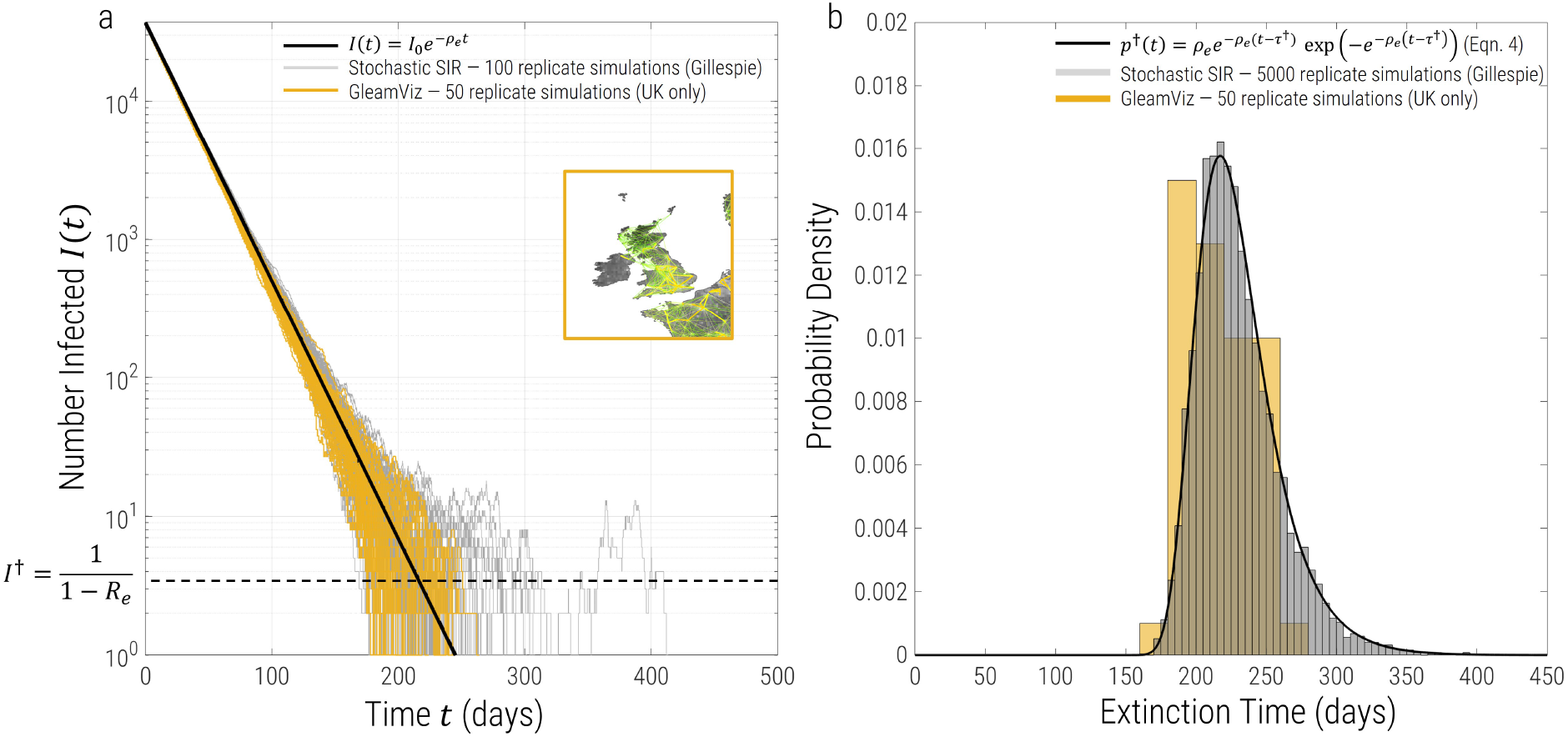
a) Simulation trajectories on log-linear scale for a decay rate of *ρ*_*e*_ = 0.043*/*day, corresponding to *R*_*e*_ = 0.7, 1*/γ* = 7 days, *I*_0_ = 3.7 × 10^4^ and an initial recovered population of *R*(0) = 6 × 10^6^. The solid black line is the deterministic prediction from Eqn.2, grey trajectories are 100 replicate Gillespie simulations of a standard SIR model, whilst the yellow trajectories are from 50 replicates using the spatial epidemic simulator GleamViz *(41, 42)* restricted to the United Kingdom with a gravity model between heterogeneous sub-populations as shown in the inset map of the UK. The dashed black line is the threshold number of infected individuals *I*^†^, below which changes in infected number of individuals is mostly stochastic. b)Probability density of extinction times for the same parameters as in a). Grey bars are a histogram of 5000 replicate simulations of Gillespie simulations normalised to give an estimate of the probability density, and the black curve is the prediction of the analytical calculation given in Eqn.3, which we see matches the simulations extremely well. The yellow bars are histograms from the GleamViz spatial epidemic simulator with 50 replicates, which we see gives similar results to the predictions of the stochastic SIR model.

### Simple random walk analysis

We can see from Fig.1a that as *I(t)* approaches extinction, as expected the trajectories become more and more varied as the number of infected becomes small. A simple heuristic treatment inspired from population genetics *(43)* defines a stochastic threshold *I*^†^, below which stochastic forces are more important than deterministic, as indicated by the dashed black line in Fig.1a; the time to extinction is then approximately the sum of the time it takes to go deterministically from *I*_0_ to 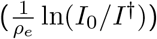 and the time it takes to go from *I*^†^ to *I* = 0 by random chance.

Assuming such a threshold *I*^†^ exists, this latter stochastic time can be approximated as follows: if there are *n* ≤ *I*^†^ infected individuals and changes are mainly random, then we are randomly drawing individuals from a pool of *n* infected individuals and *N* − *n* non-infected individuals — a binomial random walk — which when *n* ≪ *N* has standard 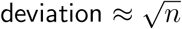 per random draw, which means we need *k* = *n* random draws, such that the standard deviation over those *k* draws is 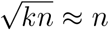; a single random draw corresponds to one infection cycle of the virus, which is *τ* = 1*/γ* days, so the time to extinction starting with *n* individuals is approximately *n/γ*.

How do we estimate *I*^†^? It is given by the threshold size at which random stochastic changes, change the number of infected by the same amount as the deterministic decline. In one cycle or generation of infection, if there was no stochasticity, the number of infected would decline by ≈ *ρ*_*e*_*I*^†^*/γ*, so equating this to the expected standard deviation of purely random changes, 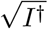, we find *I*^†^ = 1*/*(1 − *R*_*e*_), which is shown in Fig.1a for *R*_*e*_ = 0.7. Note that this threshold is closely related to Williams’ threshold theorem *(44)*, where the probability of establishment of an epidemic from a single infected individual is *p*\* = 1 − 1*/R*_*e*_, in the case that *R*_*e*_ *>* 1, which then gives a critical number of infected *I*\* ∼ 1*/p*\* = *R*_*e*_*/(R*_*e*_ − 1), below which infections changes as a random walk.

As discussed below, and in more detail in the Supplementary Materials, a more exact calculation of these considerations, using branching process theory, gives exactly the same expression for *I*^†^. This means the typical stochastic phase lasts 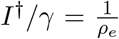 days and so adding the deterministic and stochastic phases, the mean time to extinction 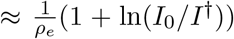 (see Eqn.4 below for a more exact expression of the mean).

### Exact branching process analysis

Given that in the regime 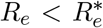, the SIR dynamics can be accurately reduced down to tracking only the dynamics of *I*, our stochastic dynamics of infections is described by a standard birth death process, where we can use standard results from branching process theory *(45, 46)* (reproduced in Supplementary Materials for completeness) that determines the probability *p*_0_*(t)* that there are exactly *I* = 0 individuals as a function of time. If *p*^†^*(t)* is the distribution of times to extinction (i.e. the probability of an extinction occuring between time *t* and *t* + d*t* is *p*^†^*(t)*d*t)*, then clearly the integral of this distribution, between time 0 and *t* is exactly *p*_0_*(t)*, and hence the distribution of times to extinction is simply the derivative of *p*_0_*(t)* with respect to time. Doing this and also taking the limit that *I*_0_ ≫ *I*^†^, we find:

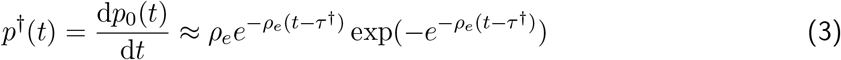

where 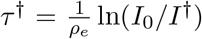, which is the time it takes for number infected to change from the initial number *I*_0_ to the critical infection size, which this calculation shows is given by 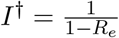; pleasingly, this is the same result as arrived by the simple random walk analysis above. Fig.1b shows a histogram (grey bars) from Gillespie simulations of the SIR model with 5000 replicates of the number of infected individuals for 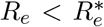. The corresponding prediction from Eqn.3 is given by the solid black line — we see that there is an excellent correspondence. In addition, Fig.3, we see that for the range of 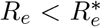 the mean extinction time from simulation fits this prediction perfectly. This result has been calculated using other methods *(38, 39)*, however, here we make clear the range of validity and the role of the stochastic threshold *I*^†^. The connection between the extinction time distribution and the extreme value Gumbel distributed *(47, 48)* arises since in a population that has *I*_0_ initially infected individuals, where each recover independently with some distribution of times, the population extinction time is the maximum extinction time of this population, which leads to extreme value statistics.

There are number of standard results for the Gumbel distribution Eqn.3, so we can write down (or directly calculate) the mean and standard deviation of the extinction time:

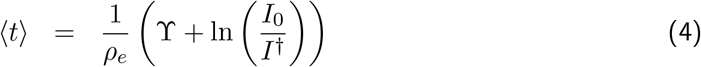

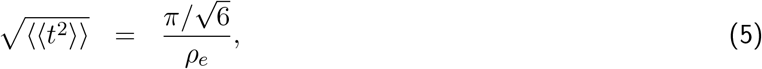

where ϒ ≈ 0.577 is the Euler-Mascheroni constant (conventionally assigned the symbol *γ*, but here *γ* is the recovery rate). We see that the heuristic calculation overpredicts the stochastic part of the extinction time by a factor of ≈ 2. Note that the standard deviation or dispersion of the distribution only depends on the inverse of the rate of decline *ρ*_*e*_ and as expected not on the initial number of infected individuals *I*_0_; hence as *ρ*_*e*_ decreases *(R*_*e*_ gets closer to 1), we see that the distribution of extinction times broadens (as we see below in Fig.4).

The cumulative distribution function for the Gumbel distribution has the well-known doubleexponential form: 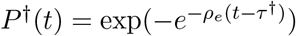, from which the inverse cumulative distribution function *T*^†^= *(P*^†^)^*−*1^ is:

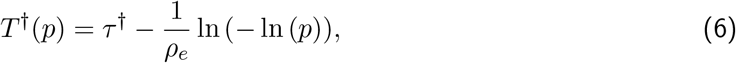

which allows direct calculation of arbitrary confidence intervals, for example, the 95% confidence intervals, by calculating *T*^†^(0.025) and *T*^†^(0.975), as well as the median 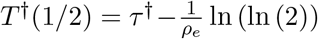 ln (ln (2)).

Finally, it is important to stress that the distribution of extinction times Eqn.3 and the following results all assume that *I*_0_ ≫ *I*^†^, so that there is a clear separation of the deterministic and stochastic phases of the decline in infections. A more general and exact result for the distribution of extinction times is given in the Supplementary Materials.

### Extinction time distribution with spatial structure and heterogeneity

A potentially valid criticism is that real populations have spatial structure and heterogeneity of contacts between regions. To make comparison to our simple predictions, we used a complex epidemic simulator GleamViz (v7.0) *(41, 42)*, which includes a gravity model of migration, where rates of migrations between sub-populations are proportional to their population sizes (see Fig.1a inset map of UK), and each sub-population based on accurate census data within a grid of 25 km. We ran 50 replicate simulations for an SIR epidemic within the United Kingdom and with zero air travel to other countries, with the same parameters as the stochastic SIR simulations in the previous section (corresponding to 12^*th*^ June 2020: *R*_*e*_ = 0.7, *γ* = 1*/*7 days, initial recovered population *R*(0) = 6 × 10^6^ – in addition, each definable sub-population in the UK was given a current infection incidence of 0.06% giving a total *I*_0_ ≈ 3.7 × 10^4^). We see the trajectories (Fig.1a – yellow lines) and histogram of extinction times (Fig.1b – yellow bars) compare very favourably to the predictions of the stochastic SIR model (black solid line and grey histogram bars); the mean and standard deviation including the gravity migration model is 211 ±16 days, which is slightly smaller than the prediction of the stochastic SIR model which has no migration or spatial structure (231 ± 30 days). This suggests that heterogeneity and migration might together have the net effect of reducing extinction times, as below we see increasing migration uniformly, has the opposite effect; nonetheless within the UK it would seem the overall effect of heterogeneity and migration is of second order to predictions of a well mixed model. Overall, we find the results of our simple model are accurate to within the width of the distributions of the extinction times.

### Accounting for population immunity: modification to theory for 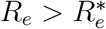

When 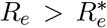, we can no longer assume that changes in the number of susceptibles has a negligible effect on *R*_*e*_ itself. In this case the decline of infections is initially non-exponential, since *R*_*e*_ decreases over time, as we see in the trajectories of Fig.2a, but at later times becomes exponential again, once *R*_*e*_ is sufficiently small that again changes in *S* become relatively negligible. As we show in the Supplementary Materials, we can use a semi-heuristic approach to calculate the steady state value of number of susceptible *S*^∞^, by integrating the SIR equations and then calculate

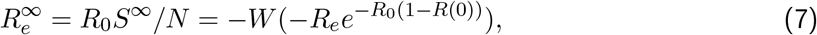

as the steady-state value of the reproductive number once infections are sufficiently small. In Fig.2a, we plot how infection would decline with 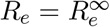, which we see has the same slope on a log-linear plot as the asymptotic mean of the simulation trajectories at later times (solid line).

**Figure 2:**
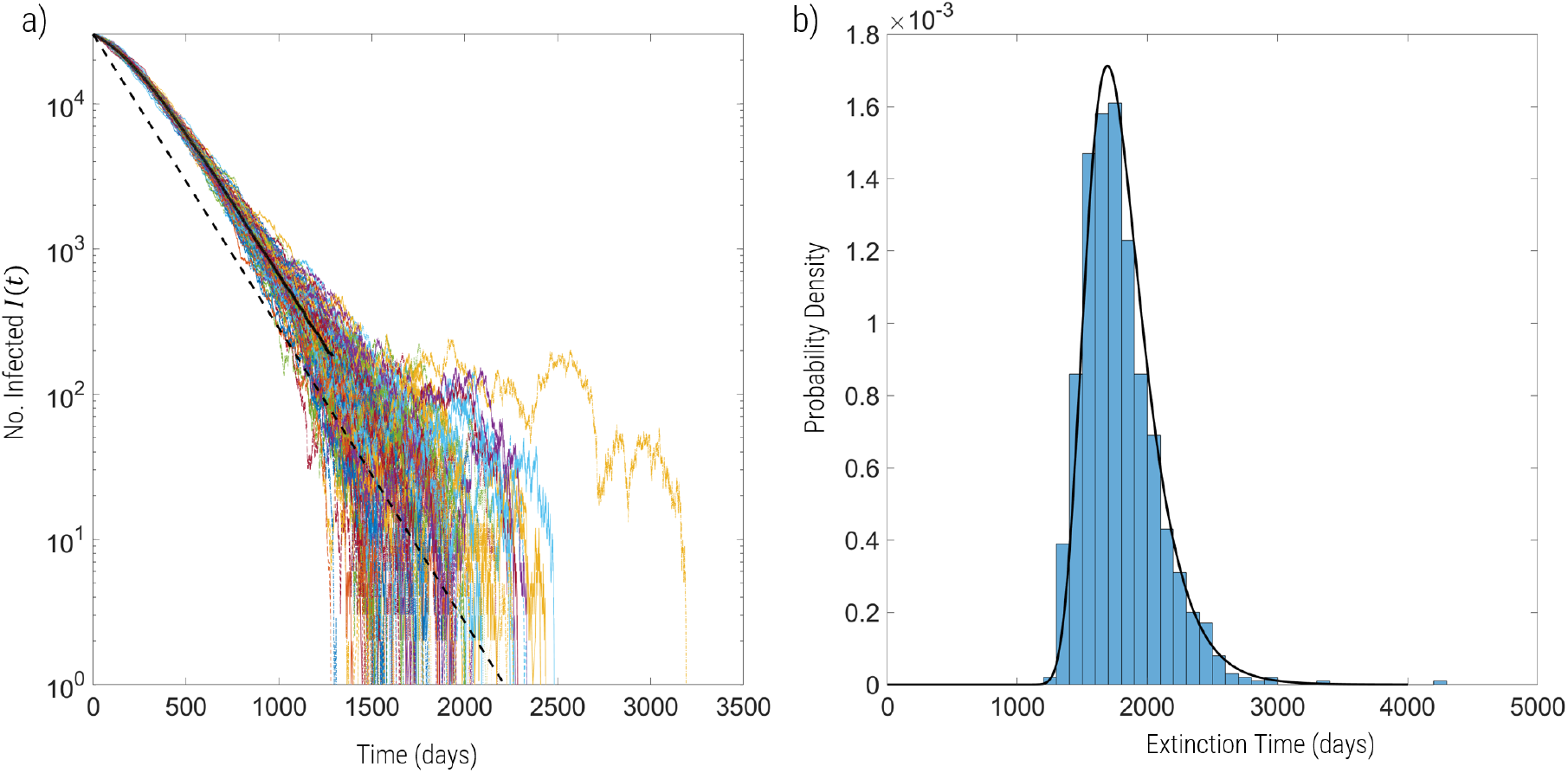
a) Trajectories of numbers infected *I(t)* for simulations with 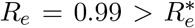 for *I*_0_ = 3 × 10^4^, *N* = 67 × 10^6^ and assuming 10% of population recovered at time *t* = 0, which gives 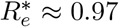; the solid line is the mean of the simulations, while the dashed line shows the decline of infections if 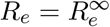 from *t* = 0, where we see the slope is the same as the mean at longer times. b) Histogram of extinction time distributions of the same simulations with prediction given by Eqn3, but with 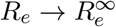 and 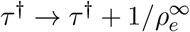, as detailed in the text.

**Figure 3:**
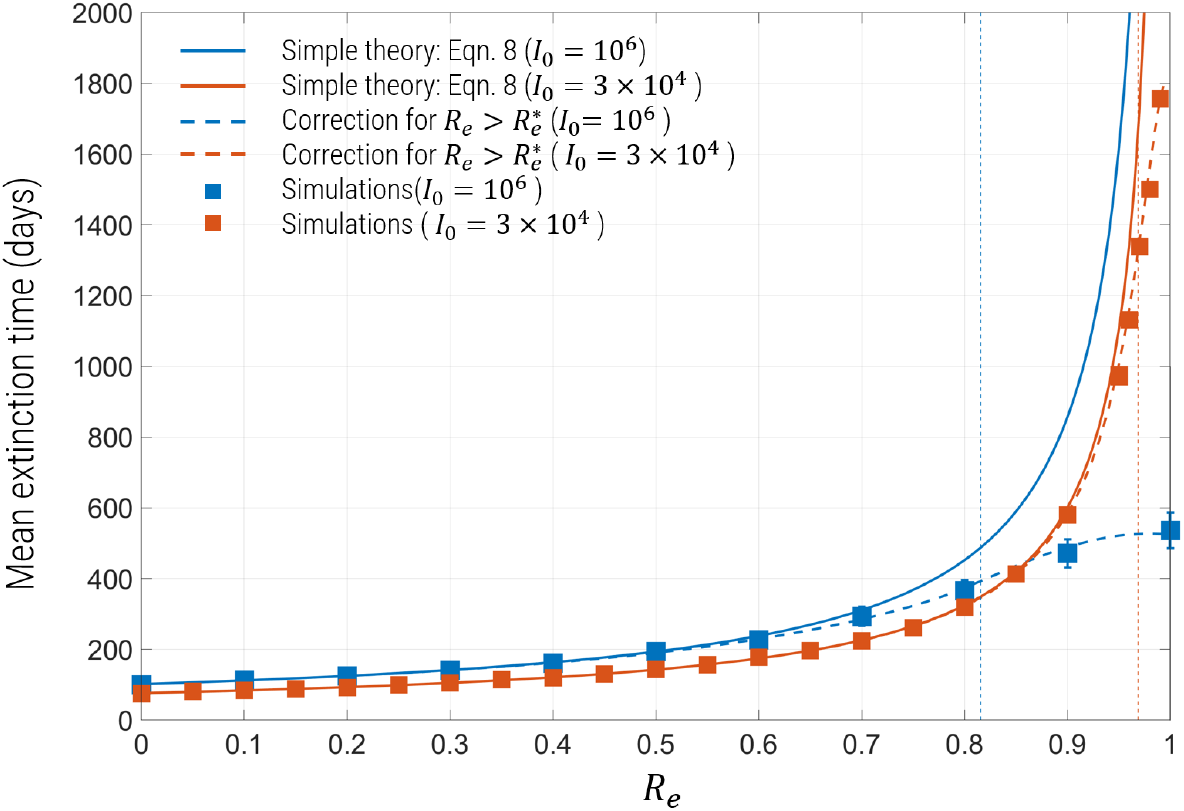
Mean extinction times for *I*_0_ = 3 × 10^4^ (red) and *I*_0_ = 10^6^ (blue) for *N* = 67 × 10^6^ and assuming 10% of population has recovered. The simple theory (Eqn.4) that assumes a constant unchanging *R*_*e*_ are shown by solid lines, while the theory modified for values of 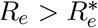 are shown as dashed lines. The values of 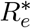 are indicated by the vertical dashed lines, which for *I*_0_ = 3 × 10^4^ is 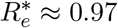 and for *I*_0_ = 10^6^ is 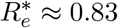. The filled squares are the mean extinction times obtained using simulations for same sets of parameters.

**Figure 4:**
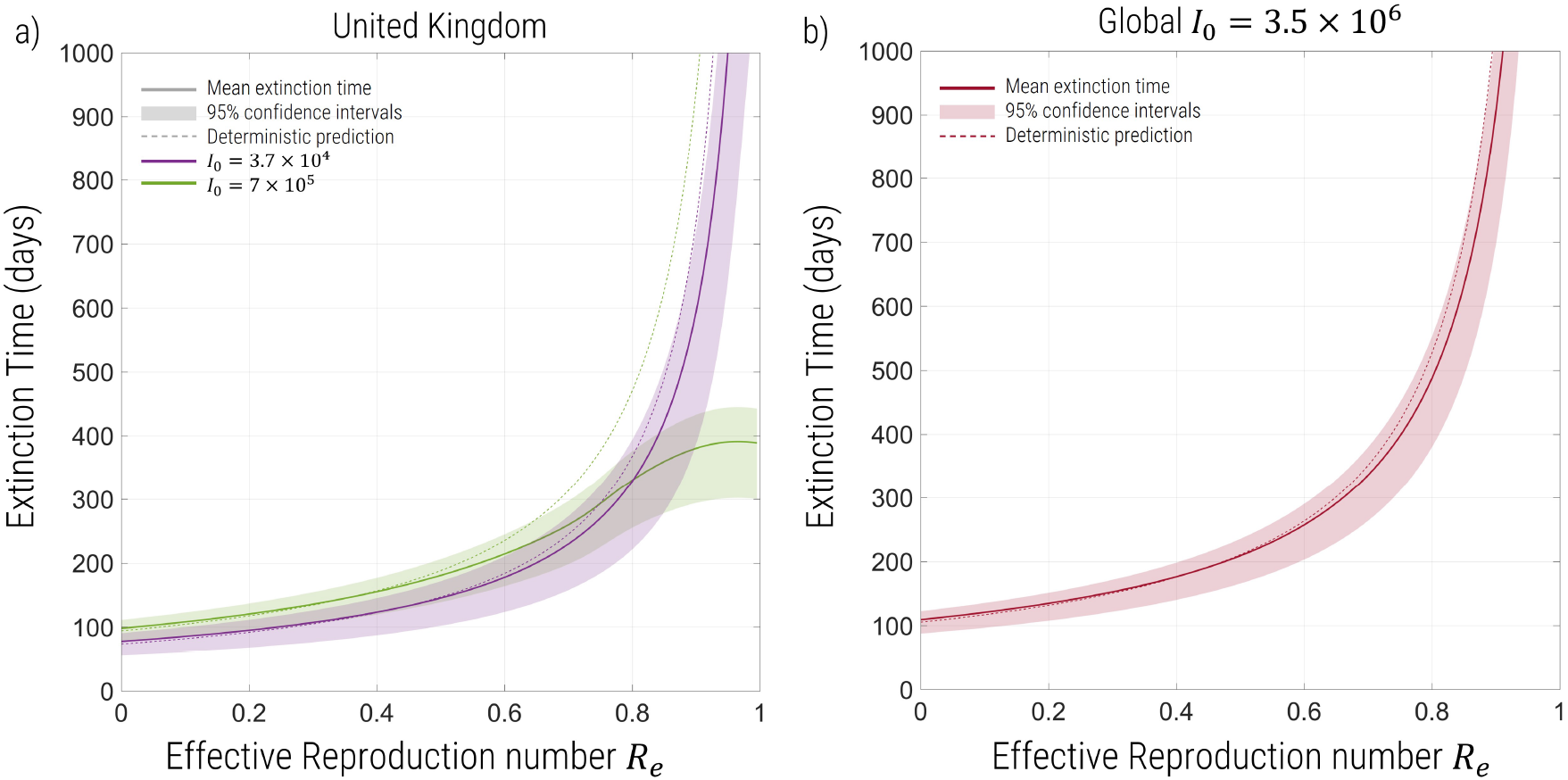
Prediction of the extinction times from analytical theory of the epidemic within the United Kingdom (a) and the pandemic globally (b) as a function of *R*_*e*_. a) For the United Kingdom we use an initial infected population of *I*_0_ = 3.7 × 10^4^ with 10% of the population recovered and immune (purple) and *I*_0_ = 7 × 10^5^ with 70% of the population immune (green), both with 1*/γ* = 7 days. b) For the global prediction, we use an initial infected of *I*_0_ = 3.5 × 10^6^ and 1*/γ* = 7 days. Solid lines are the predictions of the mean (Eqn.4), whilst each shaded region corresponds to the 95% confidence interval (Eqn.6) and the deterministic prediction (Eqn.2) are the dashed lines.

To calculate the extinction time distribution for 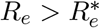, we substitute for 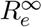 for *R*_*e*_ in Eqn.3 and in addition, make the substitution 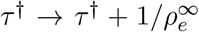, where 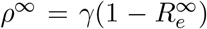 to account for the time it takes to reach this steady state. We see in Fig.2b, that this prediction for the extinction time distribution matches the histogram of times obtained by simulation very well. In addition, we see in Fig.3 that the mean extinction time of simulations fits the predictions very well for 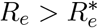. Finally, we can simply “stitch” the solutions for 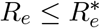 and 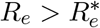, if needed, using a standard tanh switching function for *τ*^†^, as detailed in the Supplementary Materials, and shown as the dashed line in Fig.3.

It is interesting to note that for 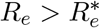, the extinction times are significantly lower for a higher number of initial infected because 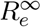 is much lower; in essence the higher infection levels lead to a significant *relative* reduction in the susceptible pool causing *R*_*e*_ to drop more dramatically.

### Extinction time predictions for SARS-Cov-2

#### United Kingdom

We first consider what this model predicts for the extinction of the SARS-Cov-2 epidemic within the United Kingdom, given an estimate of number infected of *I*_0_ ≈ 3.7 × 10^4^ with approximately 10% of the population immune for June 2020, when the epidemic was near it’s lowest incidence, and for the number of infected *I*_0_ = 7 × 10^5^ in mid-August 2021, assuming roughly 70% of the population are immune through a combination of infection and vaccination *(49)*. In Fig.4a we have plotted the estimates of mean (solid line) together with 95% confidence intervals (shaded region), given an initial number of infected *I*_0_ for various reproductive numbers *R*_*e*_ between 0 *< R*_*e*_ *<* 1. In all these estimates we assume a typical duration of 1*/γ* = 7 days for infections *(50)*.

We see three broad trends: 1) for *R >*≈ 0.6 the extinction times are very long of order years, whilst at the same time the 95% confidence intervals becomes increasingly broad (of order years themselves) meaning increasing unpredictability; 2) also for *R*_*e*_ *>*≈ 0.6, the deterministic prediction increasingly and significantly overestimates the mean extinction time; 3) for *R*_*e*_ *<* 0.5 the extinction times plateau with diminishing returns for further decreases in the reproductive number. Regarding point 3), we see that there is minimum time to extinction, given by *R*_*e*_ = 0; from Eqn.4 in the limit that *R*_*e*_ → 0, the mean time to extinction ⟨*t*⟩ → 1*/γ*(ln*(I*_0_) + ϒ), which shows the extinction process is ultimately limited by the rate of recovery *γ*, imposing a maximum speed limit on the rate of decline of infections.

In the case corresponding to summer 2020, we see that the simple stochastic SIR model predicted that extinction or elimination of the epidemic could have occurred within the United Kingdom within 4 to 5 months, which would be October/November 2020, if *R*_*e*_ can be kept to below about 0.5, and assuming no immigration of cases. More precisely, if *R*_*e*_ = 0.4, the mean time is 123 ± 15 days with 95% confidence intervals:(87, 145) days. On the other hand for *R*_*e*_ *>* 0.6 we see extinction times increase very rapidly and are of order years. However, it should be noted that should 1*/γ* = 7 days be an underestimate of the infectious period then we would expect extinction times to be approximately scaled upwards in proportion. On the other hand, taking the number of infected the following summer (mid-August 2021) where *I*_0_ ≈ 7 × 10^5^, we see extinction could have occurred within about 6 months if *R*_*e*_ *<* 0.5, or roughly by February 2022; more precisely the mean extinction time is predicted to be 157 ± 15 days (120, 177) if *R*_*e*_ = 0.4.

#### Global extinction time predictions

We can also use our calculation to make approximate predictions of global extinction times of SARS-Cov-2, with all the same broad caveats, given the simplifications of the model. We choose to examine the time to extinction from the period of summer 2020, as a hypothetical counterfactual scenario, where nations decided globally to adopt an elimination strategy. We assume in June 2020, current infections were *I*_0_ ≈ 3.5 × 10^6^, or roughly 3.5 million, based on a global death rate of 5, 000 per day. Given a global population of 7.8 billion this corresponds to a global incidence of ≈ 0.09%. We further assume that as in the UK at the time very roughly 10% of the population had acquired immunity naturally. In Fig.4b we plot how the predicted extinction time changes with effective reproductive numbers between 0 *< R*_*e*_ *<* 1 for 1*/γ* = 7 days. As would be expected, the results mirror the predictions for within the United Kingdom, except the plateau as *R*_*e*_ → 1 occurs at much longer times; for sufficiently small values of *R*_*e*_ *(R*_*e*_ *<* 0.5), the theory predicts global extinction on a timescale of slightly greater than 200 days or 7 to 8 months, whilst *R*_*e*_ *>* 0.6 lead to very long extinction times (∼years). More precisely, for *R*_*e*_ = 0.4 and assuming 1*/γ* = 7 days, we find a mean extinction time of 177 ± 15 days (95% CI: (140, 199) days). In this counterfactual scenario, global extinction of SARS-Cov-2 could have occurred by the beginning of January 2021.

## Discussion & Conclusions

Quantifying the time to extinction for elimination strategies is of great importance for learning lessons from the SARS-Cov-2 pandemic and future pandemics with similar characteristics. In this paper, we have developed a complete theory of the extinction time distribution for well-mixed stochastic SIR model, which develops two important new theoretical insights: 1) there is a stochastic phase of declining infections when *I(t)* ≪ *I*^†^ = 1*/*(1 − *R*_*e*_), where infections change only randomly and 2) there is a critical effective reproductive number 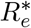— below which existing population immunity can be ignored, infections decline exponentially and standard sub-critical branching process can be applied — and above which decline is non-exponential due to the role of existing population immunity. In the latter case, we develop a heuristic approach to calculate a very accurate approximation of the distribution of extinction times. Using these results we show that for *R*_*e*_ = 0.4, extinction can be rapid and of order months with specific counterfactual predictions that from mid-June 2020, extinction would have occurred within the UK with 97.5% probability by the end of October 2020 (145 days) or globally, with 97.5% probability by the beginning of January 2021 (199 days). It is worthwhile to note that these dates are before any variants of concern had been designated, but roughly coinciding with dates at which the initial variants of concern (alpha, beta, gamma, and delta) were first detected; although, this might suggest this calculation could be invalidated, or at least more complicated, in this counterfactual scenario, the probability of the evolution of such variants would be significantly reduced with fewer global infections *(54–57)* — see further discussion below.

It is clear that with global cooperation elimination of SARS-Cov-2 was feasible on a timescale of months for the original Wuhan strain – the politics, sociology and economics of achieving such global cooperation, and the many detailed factors that contribute to controlling infections *(4–6)* is outside the scope of this paper — however, with the evolution of far more transmissible strains such as the delta and omicron-variants it is arguable that elimination becomes impractical. If we use *R*_*e*_ = 0.5 as a target, then with the original Wuhan strain estimates of the reproductive number *R*_0_ ≈ 2.5, which means assuming a completely susceptible population, the contact rate *β* would have to be reduced, using NPIs, by a 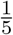, or crudely individuals would need to reduce their contacts to 20% of their pre-pandemic levels; note that in the first wave this was achieved in many countries *(6)*. For the *δ* variant, we use a representative value *R*_0_ ≈ 5 *(51, 52)* for the reproductive number, which would suggest contacts would need to be reduced to 10% of pre-pandemic levels to achieve *R*_*e*_ = 0.5, which as was seen in New Zealand was difficult to achieve, even with some level of vaccination coverage. However, for many countries with higher infection levels of the delta-variant, immunity levels are likely be higher and so this would need to be revised accordingly. Nonetheless, without vaccines that significantly cut transmission these numbers demonstrate that elimination on the timescale of months is likely only practicable early on in a pandemic like SARS-Cov-2.

Another important aspect of an elimination strategy is the role of the migration of infections between countries, which we ignore in the extinction time predictions, but the more complex simulations using GleamViz suggest play a relatively small role. The reason for this is likely the misconception that in order to enact an elimination strategy, all migration needs to be stopped. However, as long the rate of migration is not enormous, any country following an elimination strategy where *R*_*e*_ is sufficiently less than 1, there will be a stable fixed point with a small incidence of infections, due to a migration-extinction balance, and it would not be possible for a local outbreaks to occur *(53)*. Hence, at a global level as long as individual countries maintain *R*_*e*_ *<* 1, infections will decline globally even in the face of migration. It is in this context we have estimated the global time to extinction for different values of *R*_*e*_ in Fig.4b and the counterfactual scenario where extinction would have occurred by January 2021, if global elimination had been pursued from June 2020.

Elimination also potentially has the advantage of preventing the evolution of new variants of concern; although effectively anecdotal, it appears none of the variants of concern have evolved in countries pursuing elimination. This is supported by the literature on population rescue and evolution of resistance *(54–56)* which clearly predicts that resistance is more likely with increasing effective population size. In particular, recent results on the evolution of multi-site resistance *(57)* suggest the evolution of variants with changes at a large number of multiple epitope sites can happen relatively rapidly in large populations with weakly deleterious standing genetic variation.

In summary, we have demonstrated a number of new results concerning the time to extinction of an epidemic, including a complete theory for all values of the reproductive number: 0 *< R*_*e*_ *<* 1. The key contribution of this paper is to show that elimination is a viable and relatively rapid strategy, on the time scale of ∼ months not years, to eliminate infections without having to rely on herd immunity, either naturally acquired, or through vaccination; this strategy could be critical in controlling infections early on in a pandemic like SARS-Cov-2, before the development of vaccines and before the evolution of more transmissible variants, and in fact could help prevent their evolution.

## Data Availability

Code to plot extinction time predictions and distributions, as well as for performing the Gillespie simulations can be found at https://github.com/BhavKhatri/Stochastic-Extinction-Epidemic.

## Acknowledgements

I thank John McCauley (The Francis Crick Institute), Austin Burt, Vassiliki Koufopanou, Tin-Yu Hui (Imperial College) and Ace North (Oxford) for their insights and useful comments on the manuscript.

