## Supplementary materials for "A simple stochastic theory of extinction shows rapid elimination of a Sars-like pandemic"

Bhavin S. Khatri*

**This PDF file includes:**

Supplementary Text
Figs. S1
References (S1)

Supplementary Text

### Estimating extinction times from direct estimate of $\rho_e$

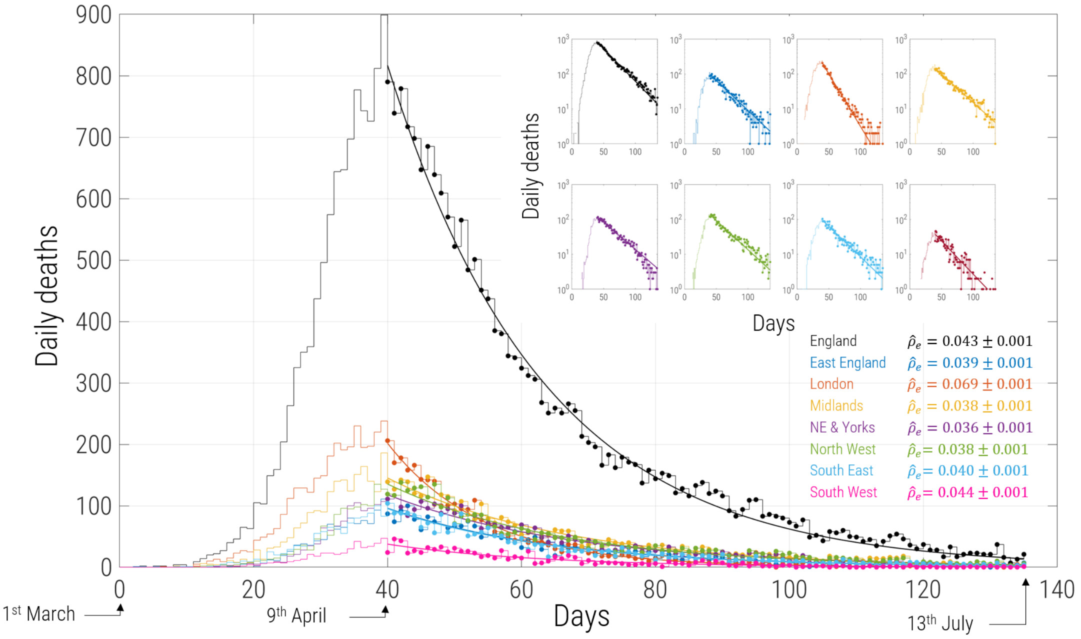


Figure S1: Data from NHS England (*S?*) on daily deaths within hospitals in England and different regions within England. Each region is fit using a simple decaying exponential from $40$ days after the 1st March 2020, where each region first shows a clear decline in number of daily deaths.

Precise predictions of extinction times based on Eqn.3 in the main text require knowledge of both $R_e$ and $\gamma$, which are generally quite difficult to estimate. However, Eqn.3 in the main text is mainly dependent on the rate of decline of the epidemic $\rho_e$, with weak dependence separately on $R_e$ and $\gamma$. $\rho_e$ can be determined more straightforwardly if we assume current daily deaths are proportional to the number infected; if infections are declining exponentially at a rate $\rho_e$ then so will the number of daily deaths, so a curve fit will give an accurate measure of $\rho_e$ even if we cannot determine the proportionality constant to translate deaths to infections. An alternative could be to look at time-series of number of daily infections per number of tests performed, to remove biases due to testing, however, here the aim is to illustrate why a direct estimate of $\rho_e$ is useful, rather than to estimate this number very accurately.

As shown in Fig.S1, fitting decaying exponentials to daily number of deaths (date of death, not date of reporting) from the NHS UK (*S?*), from the moment of decline to 13th July 2020, shows a very good fit (showing deaths are declining as a simple exponential and giving further weight to our simple model), giving an England wide estimate of $\rho_e = 0.043 \pm 0.001$ days$^{-1}$, with a range of $\rho_e = 0.036 \pm 0.001$ days$^{-1}$ (slowest decline) for North Yorkshire and $\rho_e = 0.069 \pm 0.001$ days$^{-1}$ (most rapid decline) in London. As an example, we estimate the mean time to extinction had the UK remained in lockdown from July 2020, when infections were $I_0 \approx 3.7 \times 10^4$; using the rate of decline of England, we estimate a mean extinction time in the UK as $231 \pm 30$ days ($95\%$ CI: $(187, 303)$ days), for $R_e = 0.7, 1/\gamma = 7$ days and $238 \pm 30$ days ($95\%$ CI:$(195, 310)$ days), for $R_e = 0.57, 1/\gamma = 10$ days; as we can see changing $R_e$ and $\gamma$ for a fixed $\rho_e$ does not change the predictions significantly, and we suggest in general, determining $\rho_e$ could be a more robust way to estimate extinction times.

### Branching process analysis

A birth-death process with birth rate $b = \gamma R_e$ and death rate $d = \gamma$, corresponds to a pure exponential growth ($R_e > 1$) or decay ($R_e < 1$) phase of an epidemic, when the number of susceptible individuals $S(t)$ are far in excess of the total number infected $I(t)$. In this appendix, for presentational clarity, we will use $n = I$ to represent the number of infected individuals. To write down the rate of change of the probability of $n$ infected individuals at time $t$, we need only consider the probability of having $n-1, n, n+1$ individuals and rates of transitions between them, since in the limit of infinitesimal (continuous) changes in time, we consider only changes of single individuals. The rate of transition from $n-1 \to n$ happens with rate $\gamma R_e(n-1)$ and the rate of transition from $n+1 \to n$ happens with rate $\gamma(n+1)$, which both lead to an increase in the probability of $n$, while the rate of transition from $n \to n+1$ happens with rate $\gamma R_e n$ and the rate of transition from $n \to n-1$ happens with rate $\gamma n$, which both decrease the probability of $n$. Using these facts we can write down the rate of change of the probability of $n$ at time $t$:

$$\begin{aligned}\frac{\mathrm{d}p_n(t)}{\mathrm{d}t} = \gamma\big(&R_e(n-1)p(n-1,t) \\ &- (R_e+1)np(n,t) + (n+1)p(n+1,t)\big).\end{aligned} \tag{S1}$$

However, this description isn't complete, and we need to consider how the probability of the $n = 0$ state changes, since the above equation won't work for $n-1$, since we cannot have a negative number of individuals:

$$\frac{\mathrm{d}p_{n=0}(t)}{\mathrm{d}t} = -\gamma\left(R_e np(n,t) + (n+1)p(n+1,t)\right). \tag{S2}$$

We can encompass both equations together in one by using the unit step function $U_n = 1$ for $n \geq 0$, while $U_n = 0$ for $n < 0$:

$$\begin{aligned}\frac{\mathrm{d}p(n,t)}{\mathrm{d}t} = \gamma\big(&U_{n-1}R_e(n-1)p(n-1,t) \\ &- U_n(R_e+1)np(n,t) + (n+1)p(n+1,t)\big).\end{aligned} \tag{S3}$$

For $n < 0$, as long as we have an initial condition, $p_{n<0}(t=0) = 0$, the ODEs above guarantee that $p_{n<0}(t)\forall t$. Considering each value of $n : 0 \leq n < \infty$, we have an infinite set of coupled differential equations. The standard way to solve this is to use probability generating functions:

$$G(z,t) = \sum_{n=0}^{\infty} p_n(t)z^n, \tag{S4}$$

which is in general a complex function of a complex variable $z$. Using the fact that $z\partial G(z,t)/\partial t = \sum np_n(t)z^n$, it is straightforward to show that the set of ODEs give the following first order partial differential equation for $G(z,t)$:

$$\frac{\partial G(z,t)}{\partial t} = \alpha(z)\frac{\partial G(z,t)}{\partial t}, \tag{S5}$$

where

$$\alpha(z) = \gamma(R_e z^2 - (R_e + 1)z + 1). \tag{S6}$$

This PDE can be solved by using the method of characteristics, which finds a parametric path $z(s), t(s)$ along which our original PDE is obeyed. The rate of change of $G(s)$ along this path in terms of our parameterisation is:

$$\frac{\mathrm{d}G(s)}{\mathrm{d}s} = \frac{\mathrm{d}t}{\mathrm{d}s}\frac{\partial G}{\partial t} + \frac{\mathrm{d}z}{\mathrm{d}s}\frac{\partial G}{\partial z}, \tag{S7}$$

and so with reference to the original PDE (Eqn.S5), we can identify that

$$\frac{\mathrm{d}t}{\mathrm{d}s} = 1 \quad \frac{\mathrm{d}z}{\mathrm{d}s} = -\alpha(z). \tag{S8}$$

Integrating these pair of equations gives the characteristic paths for which $\mathrm{d}G(s)/\mathrm{d}s = 0$ is a constant:

$$\frac{z-1}{z-1/R_e} e^{\gamma(R_e-1)t} = C, \tag{S9}$$

where $C$ is a constant that represents different possible initial conditions. Integrating $\mathrm{d}G(s)/\mathrm{d}s = 0$, gives

$$G(s) = \phi\left(\frac{z-1}{z-R_e^{-1}} e^{\gamma(R_e-1)t}\right), \tag{S10}$$

where $\phi$ is an arbitrary function to be determined by consideration of the initial conditions on $p_n(t)$. We can use the fact that at time $t = 0$ we assume we know the exact number of infected individuals is $n_0$ and hence, $p_n(t = 0) = \delta_{nn_0}$, where $\delta_{nn_0} = 0$ for $n \neq n_0$ and $\delta_{nn_0} = 1$ for $n = n_0$. Calculating the probability generating function for the initial delta function probability mass, we get $G(z, t = 0) = z^{n_0}$, and so we need to find a function $\phi$ satisfying:

$$G(z, 0) = \phi\left(\frac{z-1}{z-R_e^{-1}}\right) = z^{n_0}. \tag{S11}$$

Substituting $x = (z-1)/(z-1/R_e)$, we can find $\phi(x)$, to give our solution:

$$G(z,t) = \left(\frac{1 + (z-1)e^{\gamma(R_e-1)} - zR_e}{1 + R_e(z-1)e^{\gamma(R_e-1)} - zR_e}\right)^{n_0}. \tag{S12}$$

Our probability mass function $p_n(t)$, should always be normalised $\sum_n p_n(t) = G(z = 1, t) = 1$; substituting $z = 1$ we see this that the solution $G(z, t)$ behaves correctly. Finally, the reason this is all useful, is that we want to calculate the probability of zero individuals infected $p_0(t)$, which is simply given by $G(z = 0, t)$, since $0^0 = 1$:

$$p_0(t) = G(0,t) = \left(\frac{1 - e^{\gamma(R_e-1)}}{1 - R_e e^{\gamma(R_e-1)}}\right)^{n_0}. \tag{S13}$$

Substituting $n_0 = I_0$ and $\rho_e = \gamma(1 - R_e)$ gives Eqn.6 in the main text.

Differentiating Eqn.S13 to obtain the extinction time distribution, we find

$$p^\dagger(t) = \frac{\mathrm{d}p_0(t)}{\mathrm{d}t} = (1 - R_e)\rho_e n_0 \frac{(1 - e^{-\rho_e t})^{n_0 - 1}}{(1 - R_e e^{-\rho_e t})^{n_0 + 1}} e^{-\rho_e t}. \tag{S14}$$

This is an exact expression, which is valid for all values of $I_0$ and $I^\dagger$, as long as the original assumptions of the model that changes in susceptible numbers are negligible $(R_e < R_e^*)$ is true, where $R_e^*$ is given by Eqn.S19. If $I_0 \gg I^\dagger$ then we expect there to be a strong division between the deterministic phase and the stochastic phase, such that in Eqn.S14 the exponentials have sufficiently decayed such that $n_0 e^{-\rho_e t} \ll 1$, before any extinction is likely, then it is straightforward to show that the limiting form of Eqn.S14, is the Gumbel distribution, Eqn.7 in the main text, using the fact that $(1 - e^{-\rho_e t})^{n_0} \approx (1 - n_0 e^{-\rho_e t}) \approx \exp(-n_0 e^{-\rho_e t})$.

### Accounting for population immunity: modification to theory for $R_e > R_e^*$

Infections decline when $R_e < 1$. $R_e(t) = R_0 \times S(t)/N$ is in general time-dependent, composed of two factors, $R_0 = \beta/\gamma$, which for simplicity we assume is time-independent and $S(t)/N$, which will tend to decrease in time as more susceptibles become infected, so the rate of decline $\rho_e = \gamma(1 - Re)$ is in general time-dependent and increasing over time, as shown in Fig.2a, where the initial decline of infections is non-exponential. When $R_e \ll 1$, reductions in transmissions due to NPIs dominates the decrease in infections, compared to the fractional change in the susceptible pool and so $R_e \approx R_0 S_0/N$ is constant to a good approximation. However, when $R_e < 1$ but close to 1, this is no longer true, and the assumption that the number of susceptibles is roughly constant $S(t) \approx S_0$ with respect to changes in $I(t)$ and $R_e(t)$ is a poor one.

It is within this context that we would like to calculate the extinction time distribution. Although, an exact solution is not easily obtainable, we can make a semi-heuristic approximation that works very well. Initially $R_e$ is time-dependent since the changing susceptible pool has significant affect on the decline in infections. However, once infections become sufficiently small the change in the susceptible pool, per unit time, once again becomes relatively small compared to its current value and $S(t) \to S^\infty$ attains its asymptotic value $S^\infty$, at which point the constant $R_e$ assumption becomes accurate again and infections decline at a constant rate (Fig.2a). The asymptotic value $S^\infty$ cannot be calculated via standard fixed point analysis of the SIR differential equations, since the only condition for a fixed point is that $I = 0$, and this can happen for any value of $S$; the final asymptotic values depend on the initial conditions. Taking the SIR differential equations and calculating $\dot{S}/\dot{R}$ we have

$$\frac{\mathrm{d}S}{\mathrm{d}R} = -\frac{\beta S}{\gamma N} = \frac{R_0}{N} S. \tag{S15}$$

Integrating this equation, starting from an initial condition $S(0) = S_0$ and $R(0)$ to their final asymptotic values $S^\infty$ and $R^\infty = N - I^\infty - S^\infty = N - S^\infty$, then we arrive at the following transcendental equation for $S^\infty$:

$$S^\infty = S_0 e^{R_0(1 - S^\infty/N - R(0)/N)}. \tag{S16}$$

The solution can however, be expressed using the Lambert $W$ function:

$$S^\infty = -\frac{N}{R_0} W\left(-\frac{R_0 S_0}{N} e^{-R_0(1 - R(0)/N)}\right), \tag{S17}$$

where $w = W(z)$ is the solution to the transcendental equation $we^w = z$. We are interested in finding the asymptotic effective reproductive number $R_e^\infty = R_0 S^\infty/N$ in terms of the initial effective reproductive number $R_e = R_0 S_0/N$, for which the above expression can be rearranged to give

$$R_e^\infty = -W\left(-R_e e^{-R_0(1 - R(0)/N)}\right). \tag{S18}$$

We can replace $R_e \to R_e^\infty$ in Eqn.3 of the main text to calculate the distribution of extinction times to give a good approximation of the extinction times when $R_e \approx 1$ and where the above condition for constant $R_e$ is not met . However, this gives a systematic underestimate of the time to extinction, since it effectively ignores the time it takes to attain these asymptotic values, which takes of order $1/\rho_e^{\infty}$ days, where $\rho_e^{\infty} = \gamma(1 - R_e^{\infty})$. So finally an accurate and robust approximation to the extinction time distribution is obtained by the replacement $R_e \to R_e^{\infty}$ and $\tau^{\dagger} \to \tau^{\dagger} + 1/\rho_e^{\infty}$, as we can see in Fig.2b for simulations of $R_e = 0.99$ and $1/\gamma = 7$ days. Note that for sufficiently small $R_e$ the correction to $\tau$ is not needed, as $R_e^{\infty} \approx R_e$ and the assumption of constant $R_e$ is very accurate. We approximate this threshold value of $R_e$ as the value of $R_e^{\infty}(R_e \to 1)$:

$$R_e^* = R_e^{\infty}(R_e = 1) = -W(-e^{-R_0(1-R(0)/N)}). \tag{S19}$$

which is roughly the plateau value of $R^{\infty}$, which will robustly be close to the value of $R_e$ that $R_e^{\infty}$ begins to deviate from $R_e$. We can then also stitch together $\tau^{\dagger}$ for $R_e < R_e^*$ and $\tau + 1/\rho^{\infty}$ for $R_e > R^*$ using a standard $\tanh$ switching function centred on $R_e^*$ and with width $0.05$, which is used in Fig.3 and Fig.4 in the main text to provide the extinction time predictions across the whole range of $0 < R_e < 1$.

### Invariance of extinction time distribution to population sub-division

If we imagine a single population to be divided into $n$ equally sized sub-populations, each with a reproductive number $R_e$ and zero-migration between, then the extinction time distribution of $t_k$ in the $k^{th}$ sub-population will be given by Eqn.7 in the main text, but with $I_0 \to I_0/n$. Now we want to calculate the extinction time distribution of the whole population. Extinction will occur when all sub-populations have zero infected individuals. We can record the extinction times in each sub-population: $t_1, t_2, ..., t_k, ..., t_n$ and the extinction time of the whole population will be the maximum of this set: $\tilde{t} = \max\{t_1, t_2, ..., t_k, ..., t_n\}$. The cumulative distribution function of the maximum time $\tilde{t}$ will be the probability of the joint event that each sub-population $k$ has an extinction time less than $\tilde{t}$:

$$\begin{aligned} P_n(\tilde{t}) &= P(t_1 < \tilde{t}, t_2 < \tilde{t}, ..., t_k < \tilde{t}, ..., t_n < \tilde{t}) \\ &= P(t_1 < \tilde{t})P(t_2 < \tilde{t})...P(t_k < \tilde{t})...P(t_n < \tilde{t}) \\ &= (P(\tilde{t}))^n \end{aligned} \tag{S20}$$

where $P(t) = \exp(-e^{-\rho_e(t-\tau^\dagger)})$ is the CDF of the Gumbel distribution for a single population, but with $I_0 \to I_0/n$. Given the form of this equation, these calculations can be performed exactly, whereas using extreme value theory it usually required that the tails of the distribution asymptotically obey some exponential form, which allows approximate calculation. Doing these calculations we find $(P(\tilde{t}))^n = (\exp(-e^{-\rho_e(\tilde{t}-\tau_n^\dagger)}))^n$, where $\tau_n^\dagger = \frac{1}{\rho_e}\ln(I_0/nI^\dagger)$. It is then simple to show that the $n$-dependence cancels in the final result to give

$$P_n(\tilde{t}) = P(\tilde{t}) = \exp(-e^{-\rho_e(\tilde{t}-\tau^\dagger)}). \tag{S21}$$

In other words, population sub-division into equal sized isolated populations does not affect the extinction time distribution of the whole global population. In fact, it is simple to extend these arguments to any population sub-division, where $I_0 = \sum_{k=1}^{n} I_k$, where $I_k$ is the initial infected population in each, as long as the fraction of susceptible and $R_e$ is the same in each sub-population. This is not surprising, as it is just a restatement of the mean-field/well-mixed approximation that infected individuals and sub-populations all experience the same probability of encountering a susceptible individual $S_0/N$ which is set by the global number of susceptible individuals $S_0$.
